# Clinical Characteristics and Long-term Symptomology of Post-COVID-19 Olfactory and Gustatory Dysfunction

**DOI:** 10.1101/2023.02.01.23285224

**Authors:** Amelia Boyd, Dante Minichetti, Evan Lemire, Adam L Haber, Rachel E Roditi, Tanya Laidlaw, Lora Bankova

**Author notes:** These authors contributed equally (Amelia Boyd, Dante Minichetti).

## Abstract

Olfactory and gustatory dysfunction persists in 2-4% of patients who have recovered from COVID-19 beyond 6 months. Dysosmia (distorted smell) and dysgeusia (distorted taste) are frequently observed in the acute phase of many upper respiratory viral infections. However, persistent dysosmia in these patients is associated with persistent nasal inflammation. The purpose of this study was to determine the extent of patient self-assessed post-COVID-19 olfactory and gustatory dysfunction and to understand the quality and severity of the subjective symptoms over a year. A total of 426 registry participants were recruited to complete initial online questionnaires and follow up at three post-enrollment time points: 3 months, 6 months, 12 months. The Registry questionnaires assessed nasal inflammation (Sino-Nasal Outcome Test - SNOT-22), mental health (The Patient Health Questionnaire-2 -PHQ-2; Neuro-QoL Positive Affect and Well-Being - PAW-23), sleep quality (The Pittsburgh Sleep Quality Index – PSQI), In a cohort of 74 patients, clinical measurements of smell (Smell Identification Test (UPSIT) and taste (Waterless Taste Test (B-WETT)) were performed to validate self-reported measures of sensory impairment. Our data indicate that persistent COVID-19 olfactory and gustatory dysfunction is not associated with subjective measures of nasal inflammation. However, dryness of the nose/mouth, mood disturbance, and poor sleep quality are reported by the majority of participants. Participants struggle with detecting specific foul/dangerous odorants and tasting subtle flavors, which could have a negative effect on patient safety and well-being. Those infected during the earlier waves of the pandemic have more persistent and severe symptoms. Objective measure of both smell and taste are significantly reduced in the majority of participants with self-reported olfactory and gustatory dysfunction. Finally, standard anti-inflammatory topical and systemic therapy does not improve the subjective sense of smell while olfactory training is marginally more effective. This establishes persistent COVID19 olfactory and gustatory dysfunction as a chronic and difficult to treat syndrome resistant to standard anti-inflammatory therapy.

## Introduction

Among the unique symptoms of COVID-19 are the abrupt and profound loss of smell and taste. While transient disruption of normal olfaction and taste is associated with many upper respiratory infections and chronic nasal inflammatory conditions, the sudden onset and degree of severity are key distinguishing symptoms of COVID-19^1^. Importantly, up to 4% of patients who recover from COVID-19 have persistently distorted sense of smell and/or taste 6-12 months later ^2,3^.

Observational studies in patients with COVID-19 have demonstrated that up to 68% develop smell and taste disorders [1]. Of these, 55% recover within 6 months while 4% have persistent olfactory and gustatory dysfunction at the 1-year time point [2]. Many studies reported an association between olfactory epithelial expression of the SARS-CoV2 receptor, ACE2 and acute olfactory dysfunction [3, 4] Although current studies have identified mechanisms that might explain the acute loss of smell and taste due to SARS-CoV-2 ^[3-6]^, the mechanistic determinants of persistent olfactory and gustatory dysfunction remain unclear. Researchers understand that there is no indication of neuronal distress or dysfunction [3, 7, 8], which makes the unique long-term clinical experience of post-COVID-19 sensory dysfunction evermore complex and perplexing.

Few studies have yet to define the clinical predictors of persistent olfactory dysfunction and comorbidities that might distinguish patients who regain their sense of smell and taste and those who do not. A major concern for researchers was the rapid evolution of the pandemic and inability to study in isolation symptoms and characteristics specific to each strain of COVID-19 [9, 10]. Studies have attempted to examine neurological differences or mechanistic differences according to the different prominent strains of COVID-19 [8, 10]. However, researchers have yet to monitor sensory symptom and severity in relation to the 4 major waves of COVID-19.

The COVID-19 pandemic was marked by 4 major spikes in patient cases related to the introduction of 4 new strains into the population since the start of 2020. Wave 1 (February 2020 – December 2020) was the introduction on SARS-CoV-2/Alpha. Wave 2 (January 2021-May 2021 was when Beta was the predominate cause of infection, Wave 3 (June 2021 – November 2021) was dominated by Delta, and Wave 4 (December 2021 – present day) mainly consists of Omicron variants [9, 11, 12]. Although the different strains do not exist in isolation to each wave of the pandemic, the combination of contact tracing, testing, and hospitalization reports were combined to categorize these COVID strains to the 4 Waves [9, 11]. Although there is no previous work in the literature related to how different COVID strains affect sensory function, patients demonstrate great variety in acute symptomology and long-term symptomology, which suggests that potential clinical markers and indictors that can be useful to prognosis and treatments may reside in understanding the relationship of potential strain exposure and symptoms.

This study aims to investigate the clinical characteristics of post-COVID olfactory and gustatory dysfunction to identify potential predictors of persistent sensory dysfunction in patients with chronic symptomology and of self-identify recovery over the span of one year. Participants enrolled onto an online registry were sent questionnaires at 4-time intervals throughout the course of year since time of enrollment and their data was sorted according to when they were diagnosed with COVID-19 and olfactory/gustatory symptom presentation to parse out differences between the pandemic waves and patient experience. Based on the collected data, the terminology used for the sensory dysfunction experienced by participants is dysosmia: distortion of smell and dysgeusia: distortion of taste, compared to other studies that classify the experience as anosmia: loss of smell and ageusia: loss of taste [10, 13-15]. Through an online registry of 426 patients, we identified atopic disease and nasal dryness as independent predictors of prolonged duration of persistent olfactory dysfunction after COVID-19 and nuanced problems with foul/dangerous odorants and flavor recognition, which can affect patients daily physical, mental, and emotional health.

## METHODS

### Registry Recruitment and Eligibility

The study was approved by the Mass General Brigham Institutional Review Board. Participants were recruited from flyers: digitally posted on online forums for post-COVID anosmia or physically posted in specialty clinics, and advertisements on the Rally website: online platform to connect the public with research developed by Mass General Brigham Research and Massachusetts General Hospital Laboratory of Computer science. Interested participants provided informed consent and were sent electronic screening forms through REDCap® (n=587). Patients who were ≥ 18 years of age, had a history of COVID-19 (diagnosed by a physician or by positive PCR or antigen test), and experienced loss of or changes in smell and/or taste for ≥ 1 week were included in this study. Eligible patients (n=426) were electronically sent eight study questionnaires, along with questions about COVID-19 infection history and symptoms, medical history, medication use, smoking history, and demographics.

### Excluded participants

A total of 160 participants were excluded from analysis in the registry data. Participants who did not complete registry forms past the consent and screening were excluded (n=153). Participants with no history of COVID-19/sensory distortion, congenital anosmia, or anosmia due to other factors not COVID-19 were removed from further analysis (n=3). A participant record was removed for age ineligibility (n=1) and for a repeat account (n=4).

### Questionnaires

In developing the surveys, we considered our own clinical experiences working with patients with post-COVID olfactory dysfunction and designed a place where patients could self-assess their symptomology over a 12-month period since enrollment. Questionnaire installments were redistributed at 3-months, 6-months, and 12-months for participants to voluntarily complete.

Five previously validated questionnaires were included in this study: (1) Consequences of Anosmia, which assesses problems in daily life caused by olfactory dysfunction [16], (2) The Sino-Nasal Outcome Test (SNOT-22), a disease-specific quality of life survey for use in chronic rhinosinusitis [17], (3) The Patient Health Questionnaire-2 (PHQ-2), a screening scale for depression [18] (4) The Neuro-QoL Positive Affect and Well-Being (PAW-23) scale [19], which assesses overall life satisfaction and affect, and (5) The Pittsburgh Sleep Quality Index (PSQI), which assesses seven components of sleep quality and disturbances [20].

The remaining three questionnaires were designed specifically for this study: (6) Acute Anosmia, which assesses the onset, duration, and severity of dysosmia in relation to other COVID-19 symptoms and attempts to characterize dysosmia in the acute phase of COVID-19 and to rate participants’ ability to sense specific scents and flavors (7) Chronic Anosmia, which assesses chronic symptoms of dysosmia and treatments tried, and (8) Improvement survey regarding self-evaluated time and percentage of improvement from dysosmia symptoms.

### Questionnaire Scoring

Consequences of Anosmia was scored on a “yes/no” basis to give qualitative information into the daily mental, physiological, social, emotional, and safety experience for participants struggling with decreased or abnormal sense of smell. Scores were not used for diagnostic criteria or for quantitative analysis of severity for each unique consequence.

SNOT-22 domains scores were calculated for each participant: Rhinologic symptoms items #1-4, 7 (“Need to blow nose”, “Nasal blockage”, “Sneezing”, “Runny nose”, “Thick nasal discharge”), Extranasal rhinologic symptoms #5-7 (“Cough”, “Post-nasal discharge”, “Thick nasal discharge”), Ear/facial symptoms #3-8, 11 (“Ear fullness”, “Dizziness”, “Ear pain”, “Facial pain or pressure”), Sleep disturbance #13-17 (“Difficulty falling asleep”, “Wake up at night”, “Lack of a good night’s sleep”, “Wake up tired”, “Fatigue”), Mood/affective (psychological) disturbance #18-22 (“Reduced productivity”, “Reduced concentration”, “Frustrated/restless/irritable”, “Sad”, “Embarrassed”) [13]. Item #12 “Loss of smell and/or taste” was excluded from rhinology domain score and analyzed separately as it was an outlier for patients unique to the studied population compared to other items. Previous studies included items #16 (“Wake up tired”) and #17 (“Fatigue”) in the affect domain but were excluded to limit multicollinearity of mood and sleep component scores. For this study, higher SNOT-22 scores indicated a greater severity of symptoms and a lower quality of life due to persistent complications due to prior infection with COVID-19. Lower SNOT-22 scores indicated lower symptom severity to none and a greater quality of life.

PHQ-2 scores ranged from 0 to 6, where scores ≥ 3 indicate positive likelihood for depression [14]. Scores were used to determine correlations between frequency of self-assessed depressed mood and anhedonia over the past 2 weeks prior to completing the questionnaire. Scores were not used to diagnose depressive disorder or refer participants for further evaluation.

PAW-23 scores were assessed according to a T-distribution [15,17] and ranged from 23 to 107. Lower scores (<69) were considered to have increased positive affect and life satisfaction considering their loss or distortion of sensory function, while higher scores (>69) were considered to have decreased positive affect and life satisfaction considering their loss/distortion of sensory function.

Global PSQI scores ranged from 0 to 21 [16]. Seven components evaluated subjective quality of sleep, sleep latency, sleep duration, sleep efficiency, sleep disturbance, use of sleep medication, daytime dysfunction. Component scores were ranked on a scale of 0-3, indicating severity of poor sleep quality. High scores reflected greater problems with sleep than lower scores for both global and component analysis.

Acute Anosmia, Chronic Anosmia, and Improvement were used to assess qualitative experience with anosmia and dysgeusia. Acute Anosmia scores were assessed on a Likert scale to determine variety of smell/taste distortion and other symptoms, including nasal dryness, sinus pain, and phantosmia among participants. Chronic Anosmia was scored on a “yes/no” basis to see what treatments participants were trying and track specifically unpleasant/dangerous odors. Improvement questionnaire data yielded self-reported percentage scores of sensory recovery and self-determined duration of post-COVID-19 symptoms for participants enrolled in the registry.

### Clinical Measurement of Smell and Taste

Participants simultaneously enrolled in our Registry and Mediators study of post-COVID anosmia tested on UPSIT (Smell Identification Test™, Sensonics International, N.J.): a well-validated 40 multiple-choice odorant scratch-and-sniff test, and *B-WETT* (*B-WETT®-SA-27, Brief Waterless Empirical Taste Test, Sensonics International, N.J.)*: a validated 27 multiple-choice flavor strip test [18, 19]. Both tests were for one time use only. Three trained observers administered the UPSIT to 74 subjects. Participants were required to select an answer for each question even if they could not detect the odorant.

Participants were categorized by their objective olfactory and gustatory function. Participants with UPSIT scores from 0-25 were categorized as “dysosmia” (n = 24) and those with B-WETT scores from 0-16 were categorized as “dysgeusia” (n = 4) in accordance with UPSIT [18] and B-WETT [19] normative data, respectively. Participants who reported post-COVID distorted smell or taste but did not exhibit severe olfactory or gustatory dysfunction by UPSIT or B-WETT were classified as “subjective dysosmia” (sbjct dys: n = 9). Participants who scored within the “severe dysfunction” ranges on both the UPSIT and B-WETT were classified as “dysosmia and dysgeusia” (dys/dys; n = 10). Healthy controls (HC; n = 20) with no history of COVID-19 or dysosmia/dysgeusia also completed the UPSIT and B-WETT and scored within the range of normal sensory function on both tests. Three subjects reported a history of COVID-19 and transient impairment of smell and/or taste with subjective sensory recovery at the time of participation (“recovered”); interestingly, all recovered subjects scored within the normal range of UPSIT but exhibited severe gustatory dysfunction by B-WETT. Two participants with congenital anosmia also completed the UPSIT and B-WETT.

### Statistical Analysis

De-identified registry data was downloaded from REDCap® and cleaned in R. Demographic and acute COVID-19 infection features underwent contingency analysis to determine if there were differences among participants between the 4 major COVID-19 Waves.

Continuous variables such as age and BMI were analyzed through an ANOVA. State information was processed in R to count frequencies and form a heat map of the United States. Age was further processed in R between sensory dysfunction group versus recovered and analyzed using a Welch two-sample T-test. Duration plots were calculated in R using the positive covid-test date and date of self-perceived recovery and graphed in density plots using R package ‘ggridges’. Logistical regression was used to further analyze medical history and acute anosmia symptoms to create forest plots in R. Questionnaire data that produced numerical scores SNOT-22, PHQ-2, PAW-23, and PSQI were cleaned in R and plotted in Prism. SNOT-22 correlational data was generated with R package ‘corrplot’. Variables were compared using Mann-Whitney U-Test or ANOVA. Categorical parameters, Likert, and “yes/no” questionnaire data was cleaned in R and used to make Likert scales using R package ‘psych’ and ‘likert’ or uploaded to Prism to be analyzed using contingency and Chi-square. UPSIT and B-WETT scores were calculated in REDCap®. Scores were uploaded to Prism for analysis via Multiple Comparison ANOVAs.

## RESULTS

### Study population and demographics

A total of 587 participants with history of positive COVID antigen, PCR test, or physician diagnosed COVID enrolled in the online registry. Of those 426 provided answers for the questionnaires. Data from these records were used for further analysis. Participants were recruited from 42 U.S. states, with 149 (35.0%) from Massachusetts (**Fig. 1A**). Participants were pooled into clusters determined by their COVID-19 diagnosis date, according to the 4 major Waves of the pandemic: Wave 1 (n = 204), Wave 2 (n =109), Wave 3 (n = 56), Wave 4 (n = 39) (**Table 1**). For those who did not want to disclose or could not disclose their positive date were clustered in an undisclosed COVID-19 date group (n = 18) (**Table 1**). There were no notable trends in age distribution. Analysis of comorbid conditions among the participants in this Registry indicated a low incidence of chronic cardiovascular, respiratory, neurologic, or renal medical conditions with <10% of participants reporting history of hypertension, cardiac disease, lung disease, COPD/emphysema, chronic kidney disease, dementia/Alzheimer, head trauma, diabetes, stroke, or cancer (**Table 1**). Unique to our study cohort and significantly evaluated in participant across groups was nasal diseases such as nasal congestion (17.6%), chronic rhinosinusitis (8.7%), environmental allergies (42.5%). In terms of mental health, anxiety (35.2%) and depression (28.9%) were common among study participants, but the frequency was not significant (**Table 1**). These rates were higher than the reported rate in the US population.

**TABLE 1.** Demographic characteristics of Registry “Collections and analysis of clinical characterisitcs of post-COVID anosmia” participants

|  | COVID-19 Wave |  |  |  |  |  |  |  |
| --- | --- | --- | --- | --- | --- | --- | --- | --- |
| DEMOGRAPHIC CHARACTERISTICS | All Participants (n=426) | Wave 1 (n=204) | Wave 2 (n=109) | Wave 3 (n=56) | Wave 4 (n=39) | Undisclosed COVID-19 Date (n=18) | p-values | significance |
| **AGE** |  |  |  |  |  |  |  |  |
| Age, years (m ± SD) | 45 ± 14 | 45 ± 14 | 45 ± 13 | 44 ± 13 | 43 ± 14 | 48 ± 14 | 0.695 |  |
| Age 18-29, n (%) | 75 (17.6) | 34 (16.7) | 20 (18.3) | 11 (19.6) | 8 (20.5) | 2 (11.1) | 0.3595 |  |
| Age 30-39, n (%) | 95 (22.3) | 52 (25.5) | 19 (17.4) | 13 (23.2) | 8 (20.5) | 3 (16.7) |  |  |
| Age 40-49, n (%) | 101 (23.7) | 41 (20.1) | 30 (27.5) | 11 (19.6) | 12 (30.8) | 7 (38.8) |  |  |
| Age 50-59, n (%) | 87 (20.4) | 38 (18.6) | 28 (25.7) | 13 (23.2) | 7 (17.9) | 1 (5.6) |  |  |
| Age 60-69, n (%) | 60 (14.1) | 36 (17.6) | 9 (8.3) | 8 (14.4) | 3 (7.7) | 4 (22.2) |  |  |
| Age 70+, n (%) | 8 (1.9) | 3 (1.5) | 3 (2.8) | 0 (0.0) | 1 (2.6) | 1 (5.6) |  |  |
| **GENDER IDENTITY** |  |  |  |  |  |  |  |  |
| Female, n (%) | 375 (88.0) | 177 (86.8) | 103 (94.5) | 50 (89.3) | 33 (84.6) | 12 (66.7) | 0.0063 | ** |
| Male, n (%) | 50 (11.7) | 27 (13.2) | 5 (4.6) | 6 (10.7) | 6 (15.4) | 6 (33.3) |  |  |
| Non-Binary/Prefer not to say, n (%) | 1 (0.2) | 0 (0.0) | 1 (0.9) | 0 (0.0) | 0 (0.0) | 0 (0.0) |  |  |
| **RACE & ETHNICITY** |  |  |  |  |  |  |  |  |
| Non-Hispanic/Latino White, n (%) | 356 (83.6) | 174 (85.3) | 92 (84.4) | 46 (82.0) | 30 (76.9) | 14 (77.7) | 0.0149 | * |
| Hispanic/Latino, n (%) | 43 (10.1) | 15 (7.4) | 12 (11.0) | 8 (14.4) | 5 (12.8) | 3 (16.7) |  |  |
| Black/African-American, n (%) | 8 (1.9) | 6 (2.9) | 1 (0.9) | 0 (0.0) | 1 (2.6) | 0 (0.0) |  |  |
| Asian, n (%) | 11 (2.6) | 6 (2.9) | 3 (2.8) | 1 (1.7) | 1 (2.6) | 0 (0.0) |  |  |
| American Indian/Alaska Native | 1 (0.2) | 0 (0.0) | 0 (0.0) | 0 (0.0) | 1 (2.6) | 0 (0.0) |  |  |
| Another race, n (%) | 6 (1.4) | 3 (1.5) | 1 (0.9) | 1 (1.7) | 1 (2.6) | 0 (0.0) |  |  |
| Undisclosed, n (%) | 1 (0.2) | 0 (0.0) | 0 (0.0) | 0 (0.0) | 0 (0.0) | 1 (5.6) |  |  |
| **NASAL DISEASE** |  |  |  |  |  |  |  |  |
| Nasal congestion, n (%) | 75 (17.6) | 25 (12.3) | 25 (22.9) | 10 (17.9) | 7 (17.9) | 8 (44.4) | 0.0045 | ** |
| Chronic rhinosinusitis, n (%) | 37 (8.7) | 13 (6.4) | 9 (8.3) | 7 (12.5) | 0 (0.0) | 6 (33.3) | 0.0003 | *** |
| Asthma, n (%) | 69 (16.2) | 35 (17.2) | 10 (9.2) | 11 (19.6) | 10 (25.6) | 3 (16.7) | 0.128 |  |
| Atopic dermatitis or eczema, n (%) | 61 (14.3) | 26 (12.7) | 18 (16.5) | 9 (16.1) | 6 (15.4) | 2 (11.1) | 0.8835 |  |
| Immunotherapy, n (%) | 37 (8.7) | 11 (5.4) | 10 (9.2) | 7 (12.5) | 8 (20.5) | 1 (5.6) | 0.0271 | * |
| Hay fever, n (%) | 189 (44.4) | 75 (36.8) | 54 (49.5) | 26 (46.4) | 24 (61.5) | 10 (55.6) | 0.0203 | * |
| Environmental allergies, n (%) | 181 (42.5) | 73 (35.8) | 50 (45.9) | 25 (44.6) | 22 (56.4) | 11 (61.1) | 0.0401 | * |
| **UNDERLYING CONDITIONS** |  |  |  |  |  |  |  |  |
| No underlying conditions, n (%) | 144 (33.8) | 72 (35.3) | 31 (28.4) | 22 (39.3) | 15 (38.5) | 4 (22.2) | 0.4319 |  |
| Cigarette smoking, n (%) | 39 (9.2) | 20 (9.8) | 8 (7.3) | 6 (10.7) | 3 (7.7) | 2 (11.1) | 0.9271 |  |
| Hypertension, n (%) | 40 (9.4) | 22 (10.9) | 11 (2.6) | 1 (1.7) | 4 (10.3) | 2 (11.1) | 0.3506 |  |
| Cardiac disease, n (%) | 11 (2.6) | 3 (1.5) | 2 (1.8) | 2 (3.6) | 2 (5.1) | 2 (11.1) | 0.1044 |  |
| Lung disease, n (%) | 1 (0.2) | 1 (0.5) | 0 (0.0) | 0 (0.0) | 0 (0.0) | 0 (0.0) | 0.8957 |  |
| COPD/emphysema, n (%) | 4 (0.9) | 1 (0.5) | 0 (0.0) | 3 (5.4) | 0 (0.0) | 0 (0.0) | 0.0081 | ** |
| Chronic kidney disease, n (%) | 3 (0.7) | 1 (0.5) | 1 (0.9) | 0 (0.0) | 1 (2.6) | 0 (0.0) | 0.6165 |  |
| Diabetes, n (%) | 15 (3.5) | 6 (2.9) | 5 (4.6) | 1 (1.7) | 2 (5.1) | 1 (5.6) | 0.9013 |  |
| Head trauma/concussion, n (%) | 25 (5.8) | 8 (3.9) | 8 (7.3) | 5 (8.9) | 3 (7.7) | 1 (5.6) | 0.5555 |  |
| Dementia/Alzheimer's | 2 (0.5) | 1 (0.5) | 0 (0.0) | 0 (0.0) | 0 (0.0) | 1 (5.6) | 0.0274 | * |
| Stroke, n (%) | 3 (0.7) | 2 (1.0) | 0 (0.0) | 0 (0.0) | 0 (0.0) | 1 (5.6) | 0.1021 |  |
| Cancer, n (%) | 27 (6.3) | 17 (8.3) | 4 (3.7) | 2 (3.6) | 4 (10.3) | 0 (0.0) | 0.229 |  |
| Anxiety, n (%) | 150 (35.2) | 66 (32.4) | 42 (38.5) | 18 (32.1) | 16 (41.0) | 8 (44.4) | 0.6024 |  |
| Depression, n (%) | 123 (28.9) | 57 (27.9) | 38 (34.9) | 14 (25) | 10 (25.6) | 4 (22.2) | 0.5603 |  |
| PTSD, n (%) | 32 (7.5) | 12 (5.9) | 9 (8.3) | 3 (5.4) | 4 (10.3) | 4 (22.2) | 0.1223 |  |
| **BMI (CDC)** |  |  |  |  |  |  |  |  |
| BMI, kg/m<sup>2</sup> (m ± SD) | 28 ± 7 | 28 ± 7 | 29 ± 7 | 27 ± 7 | 28 ± 7 | 27 ± 7 | 0.424 |  |
| <18.5, n (%) | 14 (3.3) | 7 (3.4) | 3 (2.8) | 4 (7.1) | 1 (2.6) | 0 (0.0) | 0.5573 |  |
| 18.5 to <25, n (%) | 140 (32.9) | 63 (30.9) | 34 (31.1) | 23 (41.1) | 13 (33.3) | 7 (38.8) |  |  |
| 25 to <30, n (%) | 112 (26.3) | 52 (25.5) | 28 (25.7) | 12 (21.5) | 13 (33.3) | 7 (38.8) |  |  |
| 30 to <40, n (%) | 111 (26.1) | 64 (31.4) | 30 (27.5) | 9 (16.1) | 6 (15.4) | 2 (11.1) |  |  |
| 40+, n (%) | 25 (5.8) | 8 (3.9) | 9 (8.3) | 4 (7.1) | 3 (7.7) | 1 (5.6) |  |  |
| Undisclosed measurements, n (%) | 24 (5.6) | 10 (4.9) | 5 (4.6) | 4 (7.1) | 3 (7.7) | 1 (5.6) |  |  |

**Figure 1.**
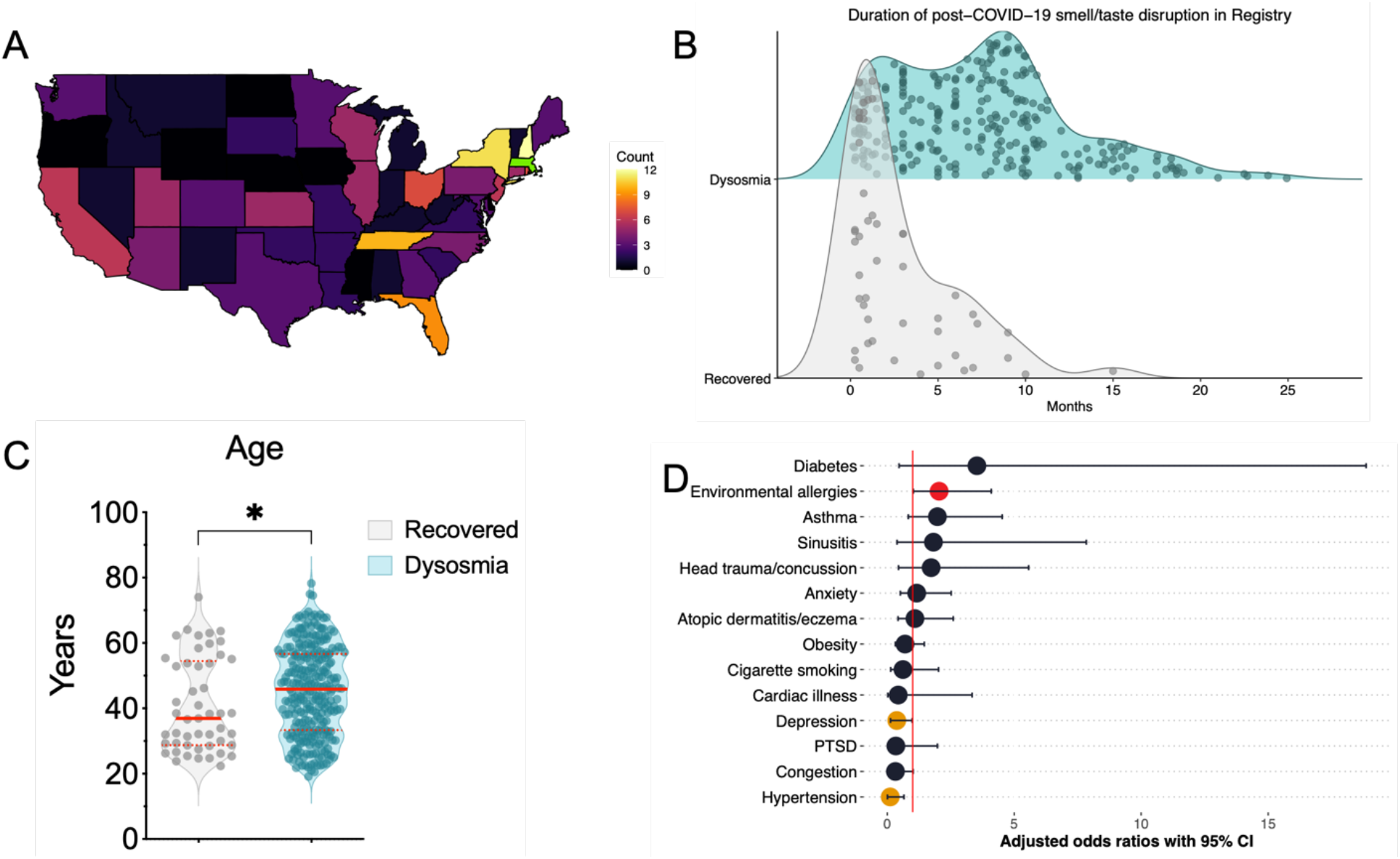
Chronic dysosmia and recovery. **(1A)** 426 participants were recruited from 42 U.S. states, including 149 (35%) from MA, 16 (3.7%) from NH, 14 (3.4%) from NY, and 13 (3%) from TN. **1B**: Recovered participants (n = 64) reported shorter duration of olfactory and/or gustatory dysfunction (μ ± SD = 2.8 ± 3 months; p = 1.45 × 10^−12^, Welch two-sample t-test) than those in the Chronic Dysosmia group (n = 362; μ ± SD = 7.4 ± 5 months). Each dot in the density plot represents one subject. Participants who reported ≥ 80% subjective improvement of olfactory dysfunction since dysosmia onset were classified as “Recovered”, and those with < 80% improvement since onset were classified as “Chronic Dysosmia”. (**1C**) Mean age in Recovered (μ ± SD = 41 ± 14 years; p = 0.044, Welch two-sample t-test) and Chronic Dysosmia group (μ ± SD = 45 ± 14 years). Each dot represents one patient. Solid lines indicate median and dotted lines indicate 1^st^ and 3^rd^ quartiles for each group. (**1D**) Forest plot of medical conditions (y-axis) in relation to recovery from post-COVID-19 dysosmia/dysgeusia. Each point denotes the adjusted odds ratio of recovery (y-axis) from multiple logistic regression, with error bars indicating 95% confidence intervals. Conditions significantly associated with recovery (p < 0.05) are shown in red and conditions approaching significance are shown in orange (p < 0.06). Conditions reported by < 5 participants were excluded from the model (stroke, lung disease). One condition (cancer) was excluded from the model due to complete separation, as all participants with a history of cancer were in the Chronic Dysosmia group. All conditions in the model had variance inflation factors < 5.

### Trajectory of post-COVID-19 olfactory and gustatory dysfunction can last over a year with no self-determined indications of improvement

Three hundred sixty-two (85%) participants reported subjective chronic post-COVID-19 olfactory and/or gustatory dysfunction for 1-24 months without significant improvement and were therefore categorized as “Chronic Dysosmia”. Sixty-four (15%) participants reported that they had regained ≥ 80% of their smell and/or taste since disease onset and were thus classified as “Recovered”. As expected, participants in the Recovered group reported significantly shorter duration of olfactory dysfunction than those in the Chronic Dysosmia group (**Fig. 1B**). Although age was not a demographically unique feature (**Table 1**), younger age (< 40 years) was related to greater likelihood of recovery (**Fig. 1C**). Interestingly, multivariable logistic regression revealed that history of environmental allergies was positively associated with recovery (p = 0.044), while depression (p = 0.055) and hypertension (p = 0.051) bordered on significance for negative association with recovery (**Fig. 1D**).

### Nasal inflammation is not an indicator of long-term symptomology of olfactory and gustatory dysfunction

To determine if clinical measures of inflammation are associated with COVID-induced anosmia and dysgeusia, we used the SNOT-22 test, validated 22-item CRS-specific quality of life instrument, demonstrated an average SNOT-22 score <20, much lower than the average reported SNOT-22 score of patients with symptomatic CRS (usually in 40-80 range) (**Fig. 2A**) ^[21, 22]^. We noted that postnasal discharge, runny nose, need to blow nose, cough, thick nasal discharge, ear fullness, facial pain/pressure, ear pain/pressure, blockage/congestion of nose, all symptoms consistent with nasal inflammation, were reported in <5% of patients with disordered sense of smell and taste after COVID19 (**Fig. 2B**). This suggested to us that persistent nasal inflammation is not associated post-COVID dysosmia or dysgeusia. When categorical symptoms rhinologic, extranasal, and ear/facial were analyzed in isolation using a Mann-Whitney U-test, there was no significant difference between groups, suggesting the factors do not contribute to the prolonged nature of the sensory dysfunction or predict recovery (**Fig. 2C**). In agreement with the limited role of inflammation, we find that only 6% of subjects in our Registry report an improvement with systemic steroids (**Fig. 2D**).

**Figure 2.**
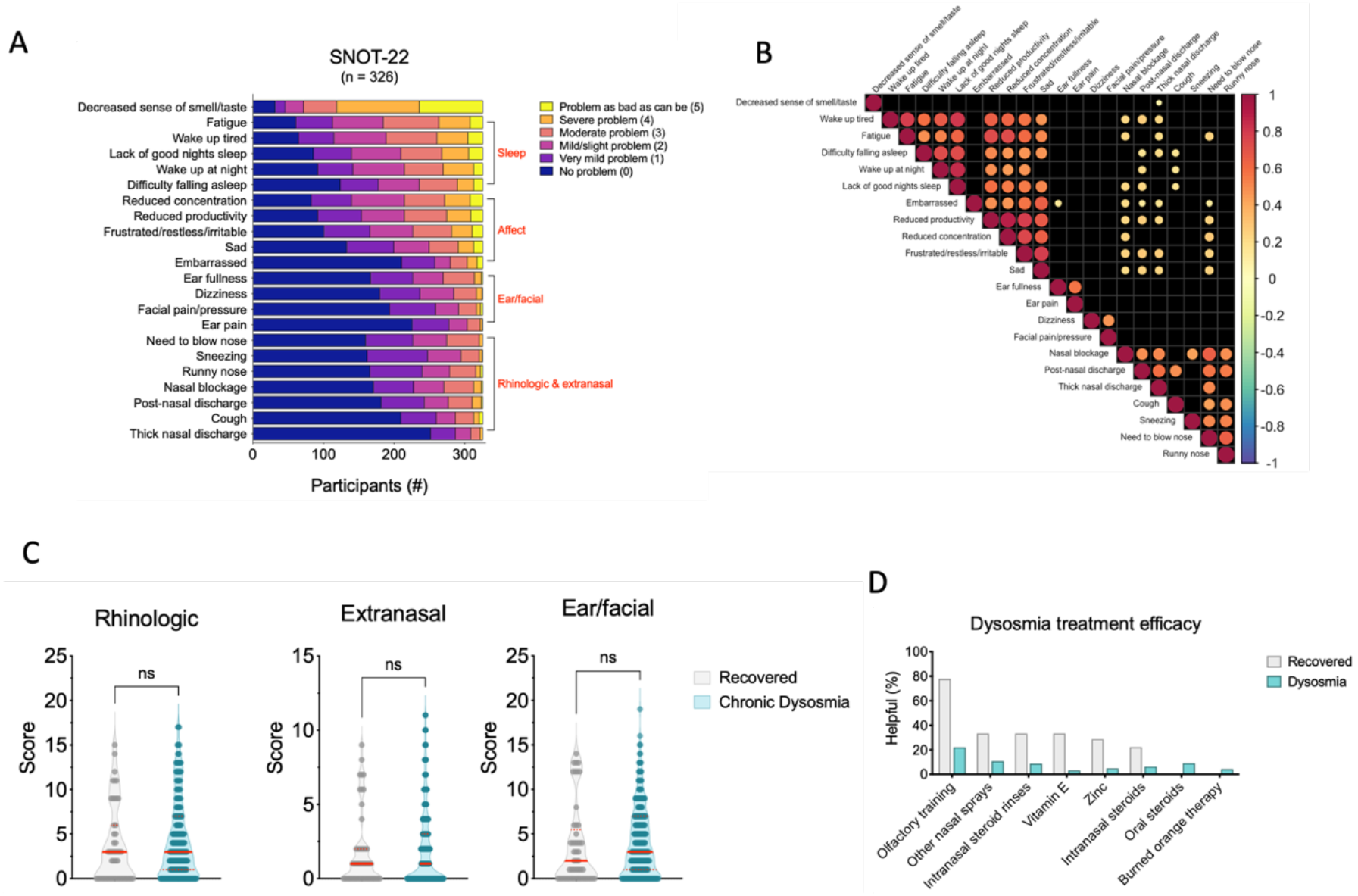
Nasal inflammation is not related to probability of recovery. (**2A**) SNOT-22 items (y-axis) grouped by domain. Bars represent counts of participants who rated each item as “Problem as bad as can be (5)” (x-axis; yellow), “Severe problem (4)” (orange), etc. (**2B**) Correlation plot of SNOT-22 items (generated with package *corrplot*). Significant associations (p < 0.05) are shown as dots arranged by hierarchical clustering (“hclust” function from R *stats* package), and insignificant relationships are shown as black squares. Dot area and color represent value of Pearson correlation coefficient. **(2C**) Violin plots of SNOT-22 rhinologic, extranasal rhinologic, and ear/facial domain scores (y-axis) for Recovered and Chronic Dysosmia groups. Each dot represents one participant. Domain scores represent the sum of scores for questions within the domain (see **Methods**). Solid lines denote median scores, and dotted lines indicate 1^st^ and 3^rd^ quartiles. Mann-Whitney U-test was used to compare domain scores across Recovered and Chronic Dysosmia groups. (**2D**) Bar graphs depicting the percent of participants (y-axis) in the Recovered (gray bars) or Chronic Dysosmia (blue bars) groups that reported subjective improvement of olfactory function after trying the denoted treatments for dysosmia (x-axis). Note: SNOT-22 rhinologic and extranasal symptoms grouped together on bar graph due to overlap (item #7, “thick nasal discharge”, appears in both domains). Item #3 (“sneezing”) appears in both rhinologic and ear/facial symptoms domains and was grouped with the ear/facial domain for the purposes of this graph.

### Poor sleep quality and affective disturbances are significant features among participants with chronic olfactory and/or gustatory dysfunction

According to SNOT-22, chronic sufferers of dysosmia and/or dysgeusia reported more problems with fatigue than self-assessed recovered participants (**Fig. 3A; Fig. 3B**). In combination with assessments of sleep quality from SNOT-22, participants were assessed on the PSQI which showed significant sleep disturbances for patients with prolonged symptoms of olfactory and gustatory dysfunction (**Fig. 3C**). Analysis of SNOT-22 categorical symptom scores also revealed that affective related issues increased in participants with greater sensory dysfunction (**Fig. 3D**). The affective domain on SNOT 22 included reports of productivity, irritability, and concentration with sad and embarrassed. Therefore, mood disturbance (sad and embarrassed) was isolated, and scores were summed to reveal that there were significantly more negative emotional repercussions to the long-term experience of olfactory and gustatory change (**Fig. 3E**). PAW-23 scores confirmed that patients with chronic dysosmia were more likely to have a negative outlook on life and their condition compared to self-perceived recovered participants (**Fig. 3F**). The results do not indicate that mood and sleep contribute to different rates of recovery; however, it is important to note the clinical features of COVID-related chronic sensory dysfunction.

**Figure 3.**
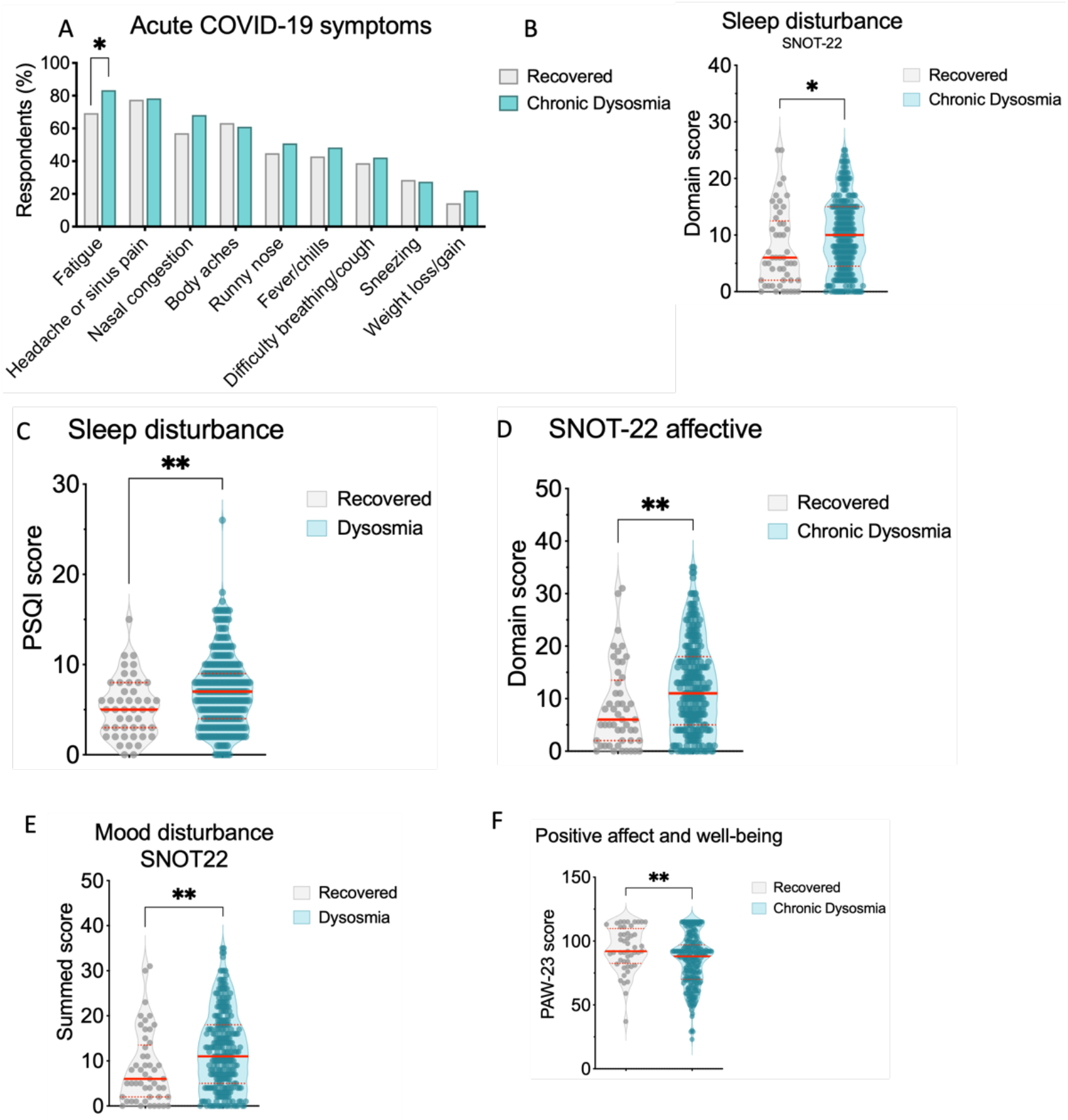
Self-assessed long-term symptoms related to chronic dysosmia are associated with poor sleep and affectivity compared to recovered participants. **(3A)** Bar graphs depicting the percent of participants (y-axis) in the Recovered (gray bars) or Chronic Dysosmia (blue bars) that experience persistent issues with the following SNOT-22 items (x-axis). (**3B**) Violin plot of SNOT-22 sleep disturbance domain (y-axis) for Recovered and Chronic Dysosmia groups (p = 0.0213). (**3C)** Violin plot of PSQI sleep scores (y-axis) for Recovered and Chronic Dysosmia groups (p = 0.0059). (**3D)** Violin plot of SNOT-22 affective domain (y-axis) for Recovered and Chronic Dysosmia groups (p = 0.0012). (**3E**) Violin plot of SNOT-22 mood disturbance isolated score of embarrassed and sad items (y-axis) for Recovered and Chronic Dysosmia groups (p = 0.0012). (**3F**) Violin plot of PAW-23 score (y-axis) for Recovered and Chronic Dysosmia groups (p = 0.0048). (**3B-F**) Each dot represents one participant. Solid lines denote median scores, and dotted lines indicate quartiles. Mann-Whitney U-test was used to compare domain scores across groups.

### Severity of olfactory and gustatory dysfunction varied depending on when participants were infected with COVID-19

Participants across the 4 major waves of COVID-19 experienced greater difficulty to distinguish between similar scents and an inability to enjoy eating (**Fig. 4A**). They also shared the experience of being able to smell faint odors that quickly vanished and the need to concentrate on what they are smelling or tasting (**Fig. 4A**). Most participants agreed and partially agreed that nose and mouth dryness was a new problem related to their post-COVID infection across the 4 Waves (**Fig. 4A**). However, Wave 1 participants were more likely to agree that food tastes different and smells are different from what they remember or once perceived prior to infection. Accounts of phantom smells also decreased across the 4 major waves, as participants responded similarly in Wave 1 and Wave 2 with more phantosmia than Wave 3 and 4 (**Fig. 4A**). Experiences of nasal pain and facial/sinus pain was not different across participant groups according to COVID Waves (**Fig. 4A**).

**Figure 4.**
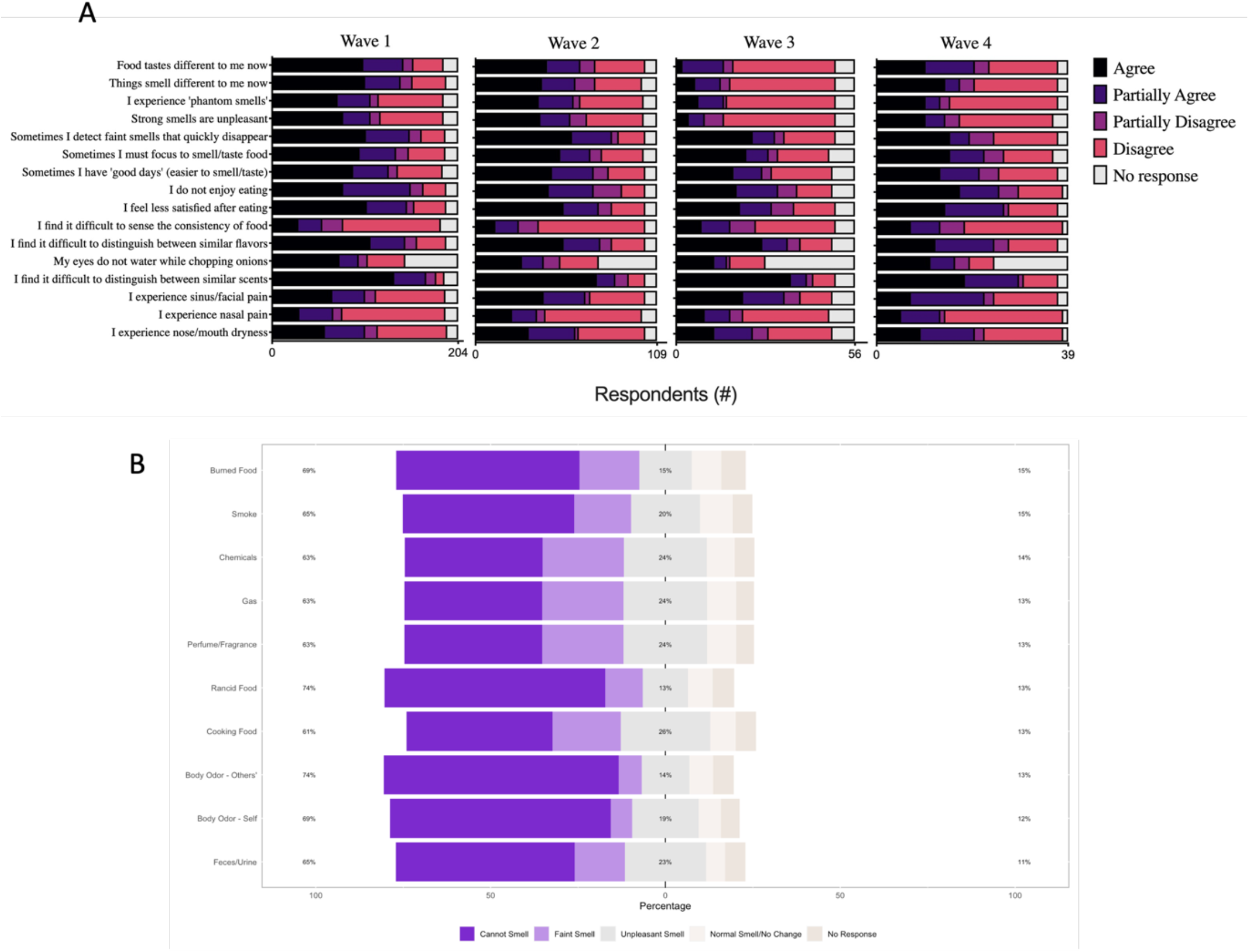
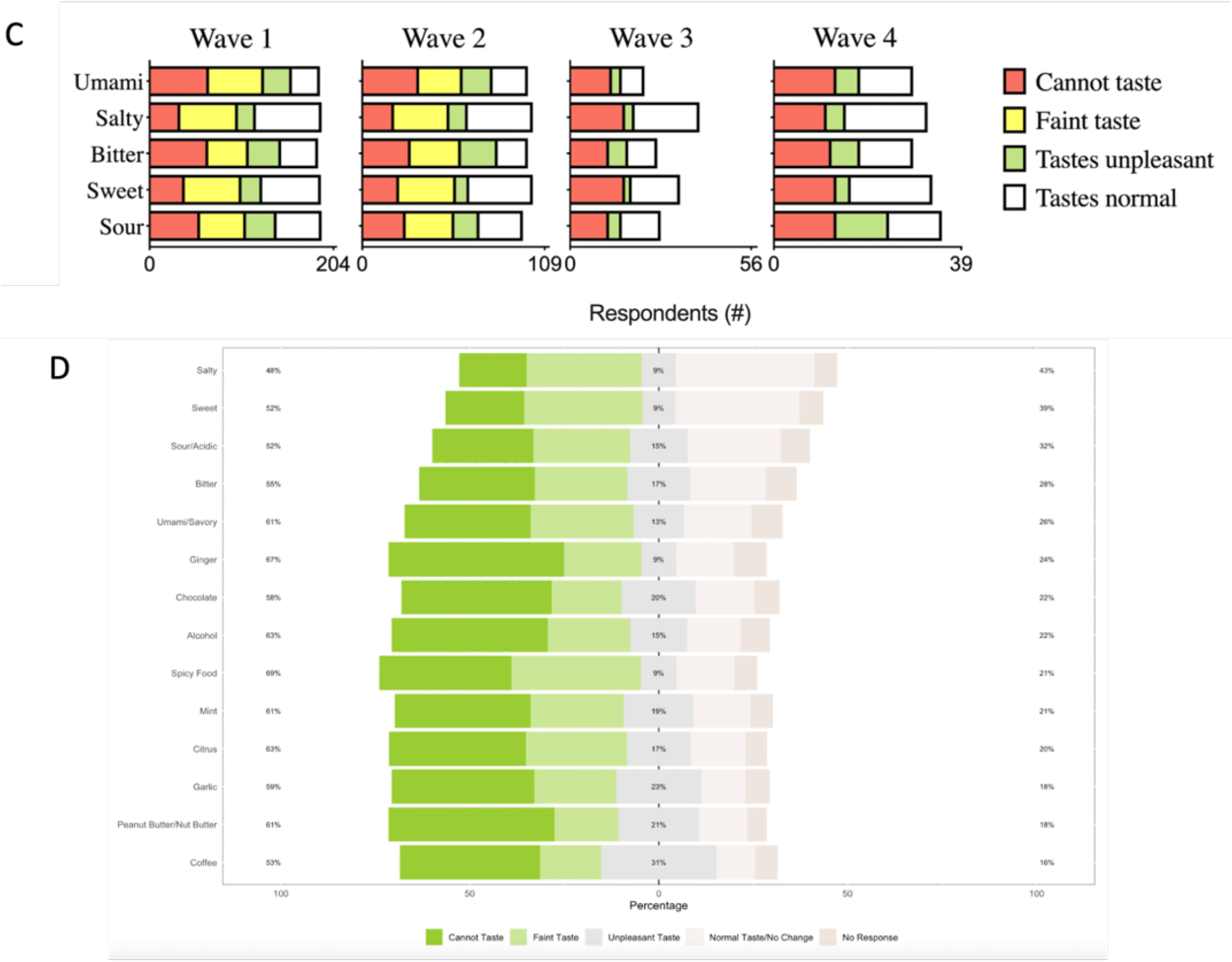
Nuanced severity of olfactory and gustatory dysfunction based on personal experience suggests a more complicated, often dangerous daily struggle with sensory impairment. **(4A)** Acute Anosmia questionnaire items appear on the y-axis. Bars represent counts of participants who rated each item on an agree/disagree Likert scale measure. Participants were isolated according to COVID-19 Wave of diagnosis and graphed separately for each wave. (**4B**) Likert graph that shows the percentage distribution of participants responses (y-axis) for specific smells (y-axis) according to a cannot smell/normal smell Likert scale. (**4C**) Acute Anosmia questionnaire items related to flavor appear on the y-axis. Bars represent counts of participants who rated each item on an agree/disagree Likert scale. Participants were isolated according to COVID-19 Wave of diagnosis and graphed separately for each wave. (**4D**) Likert graph that shows the percentage distribution of participants responses (y-axis) for specific tastes (y-axis) according to a cannot taste/normal taste Likert scale.

Participants across the 4 major waves of COVID-19 were also analyzed on their ability to taste five flavors: umami, salty, bitter, sweet, and sour. Waves 3 and 4 participants tended to not experience faint tastes (**Fig. 4C**), which is different from the shared experience of vanishing odorants (**Fig. 4A**). Overall participants struggled to detect umami and bitter across the COVID Waves (**Fig. 4C**). There were greater accounts of normal tasting function in terms of sweetness and sourness for participants in Wave 3 and 4 compared to Wave 1 and 2 (**Fig. 4C**). Saltiness appeared to become a harder flavor to detect with increasing Waves; however, the frequency of respondents that maintained normal perception of salt was high throughout the waves (**Fig. 4C**).

In terms of specific smells and particularly dangerous odors, over 70% of respondents could not detect rancid food, cooking food, or other people’s body odor (**Fig. 4B**). Participants also could no smell their own body odor (69%), burned food (69%), smoke (65%), or gas and chemicals (63%), which poses a great safety and sanitary concern (**Fig. 4B**). In a similar evaluation, participants struggled with specific favors and foods. Over 60% of respondents could not detect ginger, alcohol, mint, citrus, or nuts butters (**Fig. 4D**). The majority could not detect umami or savory (61%) out of the flavors assessed, which coincides with participant reports across the 4 COVID Waves (**Fig. 4C**). Sweetness (52%), acidity (52%), and bitter (55%) were also more commonly undetected by participants (**Fig. 4D**). However, salty flavor according to participants’ experience remained distributed on the scale between detectable to undetectable (**Fig 4D.**)

### Clinical measurements of olfactory and gustatory dysfunction cannot quantify in entirety the lived-experience and severity for patients

To determine if loss of smell and taste are independent or related symptoms of sensory dysfunction, we asked patients from our Mediators of Post-COVID Anosmia study to join our Registry and track their symptomatic progress overtime. Participants were assessed on UPSIT and B-WETT to clinically measure and evaluate sensory dysfunction to compare with self-reported measures. In accordance with the previous data, we found a variety in symptom presentation ranging in clinical severity, which created 4 phenotypes: dysosmia, subjective dysosmia (sbjct dys), dysgeusia, and dysosmia/dysgeusia (dys/dys) (**Fig. 5A**). The majority of participants in the study were patients who were diagnosed with COVID in Wave 1 and 2 (**Fig 5B**). There was not a relationship between patient type and UPSIT or B-WETT scores, which suggests that patient type is not directly dependent on the dominant COVID strain of infection for each Wave (**Fig. 5B**).

**Figure 5.**
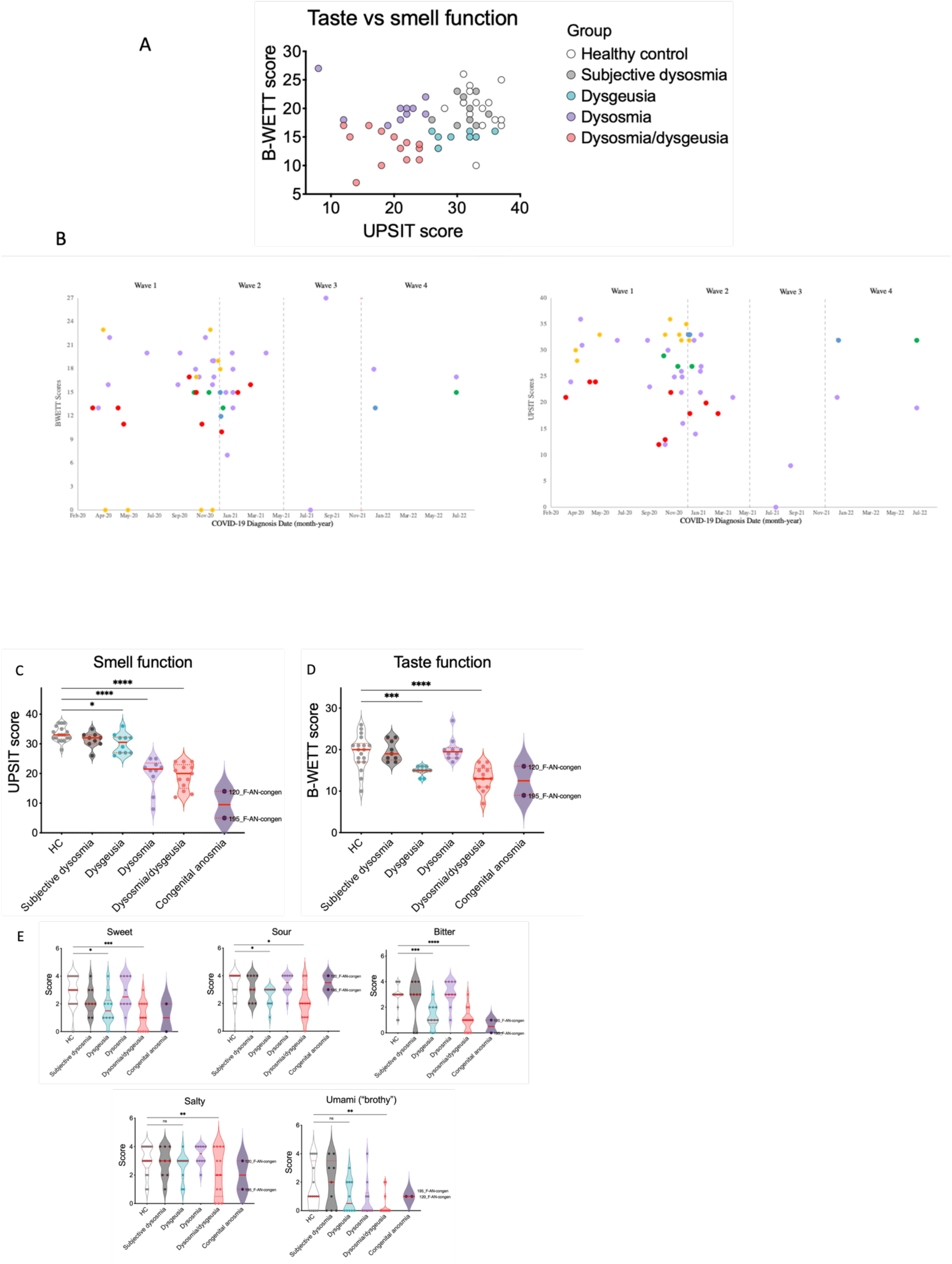
Clinical measurement of olfactory and gustatory dysfunction presented as 4 phenotypes: dysosmia, subjective dysosmia, dysgeusia, and dysosmia/dysgeusia (dys/dys). **(5A)** Scatterplot of UPSIT scores (x-axis) and B-WETT scores (y-axis) according to patient type. (**5B**) Scatterplot of B-WETT scores (left) and UPSIT scores (right) according to patient diagnosis of COVID-19 and patient type. Each dot represents one participant: purple - dysosmia, green - dysosmia, yellow – subjective dysosmia, red – dysosmia/dysgeusia, blue - recovered. (**5C**) Violin plot of UPSIT scores (y-axis) for patient type. HC vs. dysgeusia, p = 0.0214; HC vs. dysosmia, p = <0.0001; HC vs. dys/dys, p = <0.0001 (**5D**) Violin plot of B-WETT scores (y-axis) for patient type. HC vs. dysgeusia, p = 0.0009; HC vs. dys/dys, p = <0.0001. (**5E**) Violin plot of B-WETT favor sub scores for sweet, sour, bitter, salty, umami according to patient type. Sweet: HC vs. dysgeusia, p = 0.0126; HC vs. dys/dys, p = 0.0002. Sour: HC vs. dysgeusia, p = 0.0332; HC vs. dys/dys, p = 0.0124. Bitter: HC vs. dysgeusia, p = 0.0002; HC vs. dys/dys, p = <0.0001. Salty: HC vs. dys/dys, p = 0.0432. Brothy (umami): HC vs. dys/dys, p = 0.0135. (**5C-E**) Each dot represents one participant. Solid lines denote median scores, and dotted lines indicate quartiles. One-way ANOVA was used to compare domain scores across groups.

Compared to healthy controls with no history of sensory dysfunction due to COVID-19, patients with olfactory or gustatory dysfunction (dysosmia, dysgeusia, dys/dys) scored significantly lower on UPSIT, validating the olfactory distortion in participant reports (**Fig 5C**). Dysgeusia and dys/dys scored significantly lower on B-WETT compared to healthy control, validating the gustatory distortion in participant reports as well (**Fig. 5D**). In participants reports, umami and bitter flavors were the most difficult to detect, which correlates to significantly lower scores on B-WETT for dysgeusia and dys/dys versus healthy controls on bitter flavors and dys/dys versus healthy controls on umami flavor (**Fig. 4C; Fig. 5E**). According to the clinical measures, dysgeusia and dys/dys patients experienced significant difficulty perceiving sweet and sour flavors compared to healthy controls (**Fig. 5E**). Similar to self-reports, saltiness was only significantly different for dys/dys patients (**Fig. 4D; Fig. 5E**). Many patients did not struggle with saltiness compared to other flavors (**Fig. 4C; Fig. 5E**).

## DISCUSSION

The experience of post-COVID-19 olfactory and gustatory dysfunction is characterized by a wide symptom variety of quality and severity. The main goal of the study was to understand the clinical characteristics experienced by a large sample of the population of people living with post-COVID dysosmia and dysgeusia. Many existing studies have not disclosed or studied such extensive demographic features such as our Registry study [10, 12]. We wanted to focus on potential medical predispositions and threats, especially nasal disease, that could make people more susceptible to sensory dysfunction after COVID infection or provide correlational data that could predict prognosis (**Table 1**). Nasal congestion, chronic rhinosinusitis, and environmental allergies were key nasal disease criteria that demonstrated significantly different frequencies distribution across the 4 Waves of the pandemic (**Table 1)**. However, only history of environmental allergies was associated with recovery (**Fig. 1D**), suggesting a protectory effect of an overactive immune system that may have been able to minimize damage or change induced by the COVID-19 infection in the upper respiratory pathways.

COVID-19 is classified as a highly inflammatory disease that can affect both the upper and lower respiratory system [9, 11, 12]. Similar upper respiratory diseases and inflammatory infections often cause temporary olfactory and gustatory dysfunction with the presence of other illness-related symptoms. However, patients recovered from COVID-19 experience persistent sensory alteration that can last for a year post-infection with no signs of full recovery [10, 11]. The lack of subjective symptoms consistent with nasal inflammation found amongst Registry participants is remarkable considering the knowledge base surrounding such sensory phenomena during more common nasal inflammatory illnesses. SNOT-22 scores showed no difference between self-assessed recovered and participants with chronic sensory dysfunction, which suggested that recovery had no relationship with prolonged inflammatory symptoms (**Fig. 2C**). The Registry participants confirmed that nasal sprays, steroid (oral or intranasally), and rinses fail to provide temporary relief or recovery (**Fig. 2D**), making it frustrating and emotionally difficult for patients to cope with such an unprecedented trajectory of little to no progress since initial sensory dysfunction began. Participants on the Registry reported greater mood disturbances (**Fig. 3E**) and weaker positive affect (**Fig. 3F**), which can be reflected by the longevity of the post-COVID symptoms and the stigma revolving around psychologically induced symptoms.

Some studies found that patients with self-reported chronic olfactory dysfunction test objectively normal on validated scales and measures related to smell and taste [1, 23, 24]. This might suggest a perceived sensory disruption that’s out of proportion to objective findings or call to question the validity of tools that were designed for different causes of olfactory dysfunction. However, our study focused on drawing attention to the subjective, meaningful self-reported and self-monitored clinical symptoms to understand patient experience and daily struggle. Senses are based on individualized perception, which makes quantitative tests alone inaccurate for determining degrees of improvement and recovery. Based on our Registry data and clinical measures compared to healthy controls: no history of sensory dysfunction due to COVID-19 infection, and people with congenital anosmia, we found patients could fall into 4 distinct phenotypes of olfactory and/or gustatory dysfunction: dysosmia, dysgeusia, subjective dysosmia, and dysosmia/dysgeusia (**Fig. 5A**). For this study, the clinical measures were used to validate patient self-reported symptoms and compare results regarding specific smells and tastes that may influence patients’ lives on the daily. Although no relationship was found between UPSIT and specific odorants, there were major self-assessed issues with smelling foul and dangerous odors, which is concerning and problematic for quality of life and safety, reinforcing trend noted earlier in terms of mental health (**Fig. 3E; Fig. 3F**). In terms of tastes, patient reports were complementary to B-WETT scores (**Fig. 5E**) highlighting patient’s varied ability to perceive flavors and subtle differences in their food (**Fig. 4A; Fig. 5C**). As mentioned, we do not classify our patients on the anosmia or ageusia spectrum because they report extreme variation of smell and taste distortion rather than loss (**Fig. 4A**). To what extent this distortion of olfactory and/or gustatory function is based on memory or altered mechanistic pathways is unclear, and the next major goal in understanding how to help patients with chronic symptomology to regain their senses to what they were capable of prior to COVID-19.

Although SNOT-22 rhinologic and extranasal symptoms were not significantly affected among patients with chronic dysosmia/dysgeusia versus those self-identified as recovered, there were greater clinical reports of dry nose and mouth (**Fig. 2C; Fig. 4A**). The majority of participants reported lack of runny nose, nasal discharge, nasal blockage, or need to blow their nose (**Fig. 2A**), which showed no correlation to other symptoms including those related to sleep (**Fig. 2B**). Sleep quality was significantly lower for participants with chronic sensory dysfunction according to SNOT-22 (**Fig. 3B**) and PSQI scores (**Fig. 3C**). In patient fill-in reports on the PSQI, many would report restless nights due to the need to get up to drink water and snoring, which can be a result of nasal/mouth dryness. During clinical visits, patients also reported their olfactory and gustatory function decreasing through the course of the day, which could be related to dehydration and drying of the nasal cavities and mouth. The nasal lining fluid plays multiple roles: it maintains the integrity of the respiratory and olfactory epithelium, traps particulate matter and serves to transport molecules from the opening of the nares through defined trajectories along an anterior-posterior route. To reach the olfactory receptors embedded in the uppermost area of the nasal mucosa, airborne odorants, which are commonly hydrophobic molecules, are transported through the aqueous nasal mucus. Components of the nasal lining fluid that contribute to olfactory function include the odorant binding proteins and enzymes involved in the biotransformation of odorants. Further mechanistic studies might uncover the link between nasal dryness and disrupted sense of smell and taste after COVID19.

A limitation to the study was the lack of gender and racial/ethnical diversity (**Table 1**); however, the Registry was provided in English and Spanish. Participants who needed another language could request for assistance and/or translation. Future work comparing clinical characteristics to mechanistic data of post-COVID olfactory and gustatory dysfunction will help to reveal difference among patients infected with COVID at different timepoints within the pandemic, which can lead to better treatments and better outcomes for patient’s mental and physical recovery.

## CONCLUSION

Persistent olfactory and gustatory dysfunction is a distinctive feature of COVID-19 that develops with acute symptoms of infection but does not ameliorate when other symptoms such as congestion, inflammation, sore throat, cough, or fever recover after the course of illness. Researchers have not found traces or remnants of the COVID virus in the epithelium of the nasal passage that could suggest prolonged infection or cyclic reinfection. Through this longitudinal study, we were able to gain insight into the quality and severity of olfactory and gustatory dysfunction that patients are still experiencing post-infection and illness recovery. From the registry, we found that participants with chronic dysosmia and/or dysgeusia experience more sleep problems and more negative affect than recovered patients. We discovered a unique relationship between diagnosis date and COVID-19 Wave that indicated some subtle difference in patient experience between ability to smell and taste and correlate more positive prognosis for participants more likely infected with Delta and Omicron in later Waves 3 and 4 rather than participants more likely with SARS/Alpha or Beta in earlier Waves 1 and 2. Clinical measures from our mechanistic study allowed us to validate self-reported data and demonstrate how unique features of post-COVID-19 olfactory and/or gustatory dysfunction cannot be measured or shown through such validated tests. We determined significant clinical characteristics related to nasal/mouth dryness and that could be contributing to the long-term sensory dysfunction in participants. Although the study relied on self-reported data and we could not get all enrolled Participants to be clinically measured on their smell and taste abilities, there was profound similarity among the participants recruited that point to new avenues of possible research that can further expand the field of expertise related to the COVID-19 pandemic and persistent sensory dysfunction due to disease. Our next goal is to understand the mechanism related to these clinical features in our current ongoing study and compare them with our Registry data to develop prognosis trajectories for patients according to potential COVID-19 strain exposure and testing more targeted courses of treatment.

## Data Availability

All data produced in the present study are available upon reasonable request to the authors

## Notes

### Competing Interest Statement

The authors have declared no competing interest.

### Funding Statement

This study was funded by institutional funds of the Brigham and Women's Hospital Division of Allergy and Clinical Immunology.

### Author Declarations

The study was approved by the Mass General Brigham Institutional Review Board.

